# Estimating enteric fever seroincidence in rural western Cambodia: findings from a population-based cross-sectional serosurvey

**DOI:** 10.64898/2026.02.01.26343908

**Authors:** Meiwen Zhang, Natnaree Saiprom, Rupam Tripura, Lek Dysoley, Phal Chanpheakdey, Vanna Moul, Arjun Chandna, Elizabeth M Batty, Sue J Lee, Richard J Maude, Nicholas PJ Day, Thomas J Peto, Narisara Chantratita, Richelle Charles, Yoel Lubell, Kristen Aiemjoy

## Abstract

**Introduction:** Enteric fever, caused by *Salmonella enterica* serovars Typhi and Paratyphi, remains an important cause of febrile illness in low- and middle-income countries (LMIC). However, the exact burden is difficult to estimate due to limitations in diagnosis and surveillance.

**Methods:** Samples from a representative cross-sectional household serosurvey in rural western Cambodia were used to estimate enteric fever seroincidence among children and young adults. Participants were enrolled between April 7 and December 10, 2023, and 529 samples from individuals aged 5-25 years were analysed. Using IgA and IgG responses to hemolysin E antigen and established models of antibody decay after infection, we estimated seroincidence, conveying the rate of new infection in the population. Participants were enrolled between April and December 2023, and 529 samples from individuals aged 5-25 years were analysed.

**Results:** The overall enteric fever seroincidence rate was 161.8/1,000 person-years (95%CI: 144.6, 181.0). Among children 5-15 years, it ranged between 149.5 (95%CI: 128.7, 173.7) and 239.0 (95%CI:169.2, 337.6) across districts. Seroincidence was numerically higher among children 5-15 years from households with unimproved drinking water sources or sanitation facilities, and districts with a higher proportion of households with unimproved WASH.

**Conclusion:** These findings potentially support the introduction of the typhoid conjugate vaccine in rural western Cambodia. However, additional data on the relative contribution of S. Typhi versus S. Paratyphi is needed. We also demonstrate seroincidence as an adjunctive valuable surveillance tool in LMICs where facility-based surveillance may be inadequate.

**Key questions:** *What is already known on this topic:*

- Cambodia is estimated to have a high burden of enteric fever with a modelled incidence of 1.1/1,000 person-years among those under 20 years in 2023, but lacks population-level data.
- Existing estimates likely underestimate the burden because facility-based surveillance is affected by limitations on diagnosis sensitivity and healthcare access, and the modelled approach lacks country-specific incidence data.
- Validated antibody kinetics models using Hemolysin E (HlyE) antigen of *Salmonella* serovars Typhi and Paratyphi enable estimation of enteric fever seroincidence from cross-sectional serosurveys.

*What this study adds:*

- This study provides the first population-based estimates of enteric fever seroincidence in rural western Cambodia using HlyE antibody kinetics.
- Children 5-15 years showed high enteric fever seroincidence that potentially meets the WHO definition for a high-typhoid-incidence setting.
- Seroincidence varied geographically and by age, with higher seroincidence in districts with poor water, sanitation and hygiene (WASH) access, and the patterns suggest variations in *S*. Typhi and *S*. Paratyphi attribution.

*How this study might affect research, practice or policy:*

- Our findings indicate a substantially higher enteric fever burden in rural western Cambodia than previously reported.
- High seroincidence among children supports the consideration of the typhoid conjugate vaccine introduction, while highlighting the need for blood culture data to clarify *S*. Typhi and *S*. Paratyphi attribution to guide vaccination strategies better.
- Improving WASH access should also be prioritised to reduce transmission of both *S*. Typhi and *S*. Paratyphi.
- Seroincidence estimation from cross-sectional serosurveys offers a scalable approach for settings where blood culture surveillance is limited.

## Introduction

Enteric fever, caused by *Salmonella enterica* serovars Typhi and Paratyphi A, B, or C, remains common in many low and middle-income countries (LMICs), especially in settings with poor water and sanitation infrastructures (WASH).^1^ Globally, there were an estimated eight million cases in 2023, resulting in more than 80,000 deaths, two-thirds of which were among children under 15 years.^2^ Enteric fever incidence is typically estimated via blood culture-based clinical surveillance, but this method has limitations. Sensitivity is affected by blood volume and prior antibiotic use, and diagnostic services are often limited or costly, especially in rural LMIC settings.^3^ Consequently, traditional surveillance methods capture only a fraction of cases, particularly missing those who do not seek care in formal health facilities or live outside urban areas.

The WHO recommends the typhoid conjugate vaccine (TCV) use in settings with endemic or drug-resistant typhoid to reduce morbidity, mortality, and unnecessary antimicrobial use.^4,5^ Vaccination campaigns targeting children 15 years or younger in LMICs are advised for maximum impact. However, due to the aforementioned challenges, many countries, including Cambodia, lack population-level data to support vaccine introduction. Such data, including geographic and demographic variations in the disease burden, are important to direct immunisation programmes towards the populations with the greatest needs and achieve maximum impact.

As part of the South and Southeast Asian Community-based Trials Network (SEACTN),^6^ a cross-sectional household survey was conducted in rural western Cambodia to estimate the seroprevalence of pathogens causing fever.^7^ Within this survey, we also estimated seroincidence of enteric fever based on antibody responses to the Hemolysin E (HlyE) antigen of *Salmonella enterica* serovars Typhi and Paratyphi using established models of HlyE-specific antibody kinetics from blood culture-confirmed *S*. Typhi and *S*. Paratyphi cases.^8^

## Methods

### Study design

The SEACTN household survey study protocol, including details on the study setting, population, sample size, sampling methods, and procedures, has been published previously.^7^ In brief, this cross-sectional survey was conducted in one of the operational areas of SEACTN, across 82 villages in four districts within Battambang and Pailin provinces, western Cambodia. The sample size was designed to estimate the seroprevalence of multiple febrile pathogens and the prevalence of selected non-communicable diseases at the site level.

### Patient and Public Involvement statement

Patient and public involvement were described in the published study protocol.^7^ In summary, community members contributed to questionnaire validation and community mobilisation activities were conducted prior to data collection. Study findings are being shared with national stakeholders to inform enteric fever control and vaccination policy discussions in Cambodia.

### Participants

Villages and households were randomly selected, and all household members were invited to participate to obtain site-representative samples. Participants were recruited between April 7, 2023 and December 10, 2023. For this enteric fever sub-study, we included participants aged 5 to 25 years with valid HlyE IgA and IgG measurements, based on the age range supported by available antibody decay kinetics models and sample integrity.^8^

### Procedures

Household characteristics, including main drinking water source, sanitation facilities, and asset-based household wealth status, were collected through questionnaire interviews with the household head.^7^ Types of drinking water source and sanitation facilities (WASH) were categorised as ‘improved’ or ‘unimproved’, with an additional ‘no facility/ open defecate’ category for sanitation facilities. Improved WASH were defined as those that protect the water source from contamination and separate human waste to avoid human contact to ensure hygiene^9^. Households with both improved water and sanitation facilities were categorised as ‘Both improved’, and all others as ‘≥1 unimproved’. Household wealth status was classified as ‘Poor’, ‘Middle’ or ‘Rich’ using EquityTool, which applies a subset of Cambodia-specific wealth index questions from the 2014 Demographic and Health Survey to position household wealth relative to national distributions.^10^

Venous samples were collected and centrifuged in the field laboratory. Aliquots were shipped to and tested in the central laboratory at the Mahidol Oxford Tropical Medicine Research Unit in Bangkok, Thailand.^7^ Aliquots were stored at -20°C or below without freeze-thaw until analysis. Serum samples from participants aged 5-25 years were tested with a kinetic ELISA to quantify IgA and IgG responses to HlyE of *Salmonella enterica* serovars Typhi and Paratyphi as described by Aiemjoy and colleagues.^8^

We obtained written informed consent from all eligible participants and the parents or guardians of participants younger than 18 years before the questionnaire interview and collection of blood samples. We also obtained written assent from children aged 15-17 years. This study protocol was approved by the Oxford Tropical Research Ethics Committee (OxTREC Ref: 6-22) and the Cambodian National Ethics Committee for Health Research (December 23, 2022 NECHR). This completed study is registered on clinicaltrials.gov (NCT05389540).

### Statistical analysis

Age-dependent antibody responses were modelled using generalised additive models (GAMs) with penalised splines for age.^11^ District-specific smooths were fitted to allow non-linear age trends to vary by district. Model-based predictions with 95% confidence intervals (CIs) were generated.

Seroincidence was estimated using maximum likelihood profiles of our participants’ quantitative HlyE IgA and IgG ELISA measurements fitted to the established within-host two-phase antibody kinetics models, assuming Poisson-distributed time between sample collection and infections.^8^ These antibody kinetics models, developed from blood culture-confirmed cases and demonstrating similar antibody decay rates across countries, account for individual variability and laboratory-specific measurement noise, supporting their applicability to our study population. Standard errors were adjusted using cluster-robust variance estimation at the village level to account for within-cluster correlation induced by the cluster sampling design. Seroincidence estimates presented in the main results were derived using both IgA and IgG responses, whereas estimates based on single antibody isotypes are provided in Supplementary Table 1.

The seroincidence estimates were calculated using the serocalculator R package.^12^ IgA and IgG responses to HlyE from our study participants were input into the serocalculator, stratified by age groups (5-15 years, 16-25 years), geographic district (Pailin, Koas Krala, Rukhakiri, Samlot), household WASH categories, and wealth status to explore variations in transmission pattern. Age stratification reflected the target population for TCV introduction (≤15 years). To assess the potential impact of seasonal variation and timing of sample collection on seroincidence estimates, we conducted a sensitivity analysis stratifying seroincidence by month of data collection using the same analytical approach. Differences in seroincidence between groups were assessed using a two-sample z test, and statistical significance was defined as p-values <0.05. Seroincidence estimates of enteric fever (*S*. Typhi and *S*. Paratyphi) were reported per 1,000 person-years with 95% CIs. To improve statistical robustness, seroincidence estimates are not calculated for strata with fewer than 10 observations due to limited reliability. All analysis was done using R statistical software version 4.3.3.

## Results

Among 613 eligible individuals aged 5-25 years, 563 (91.8%) participated. Of these, 529 participants (94.0%) from 273 households provided blood samples with valid laboratory results and were included in the analysis (Figure 1). The median age was 12 (IQR: 8-17) years (Table 1). Age distribution was similar across districts. Compared to Palin and Samlout, a higher proportion of participants from Koas Krala and Rukhakiri districts lived in households with ≥1 unimproved WASH (Koas Krala: 85·3%, n=116 participants; Rukhakiri: 85·2%, n=46; Pailin: 47·5%, n= 67; Samlout: 58·6%, n=116), and higher proportion from the “Poor” households (Koas Krala: 25·0%, n=34; Rukhakiri: 33·3%, n=18; Pailin: 12·8%, n= 18; Samlout: 9·1%, n=18).

**Table 1.**
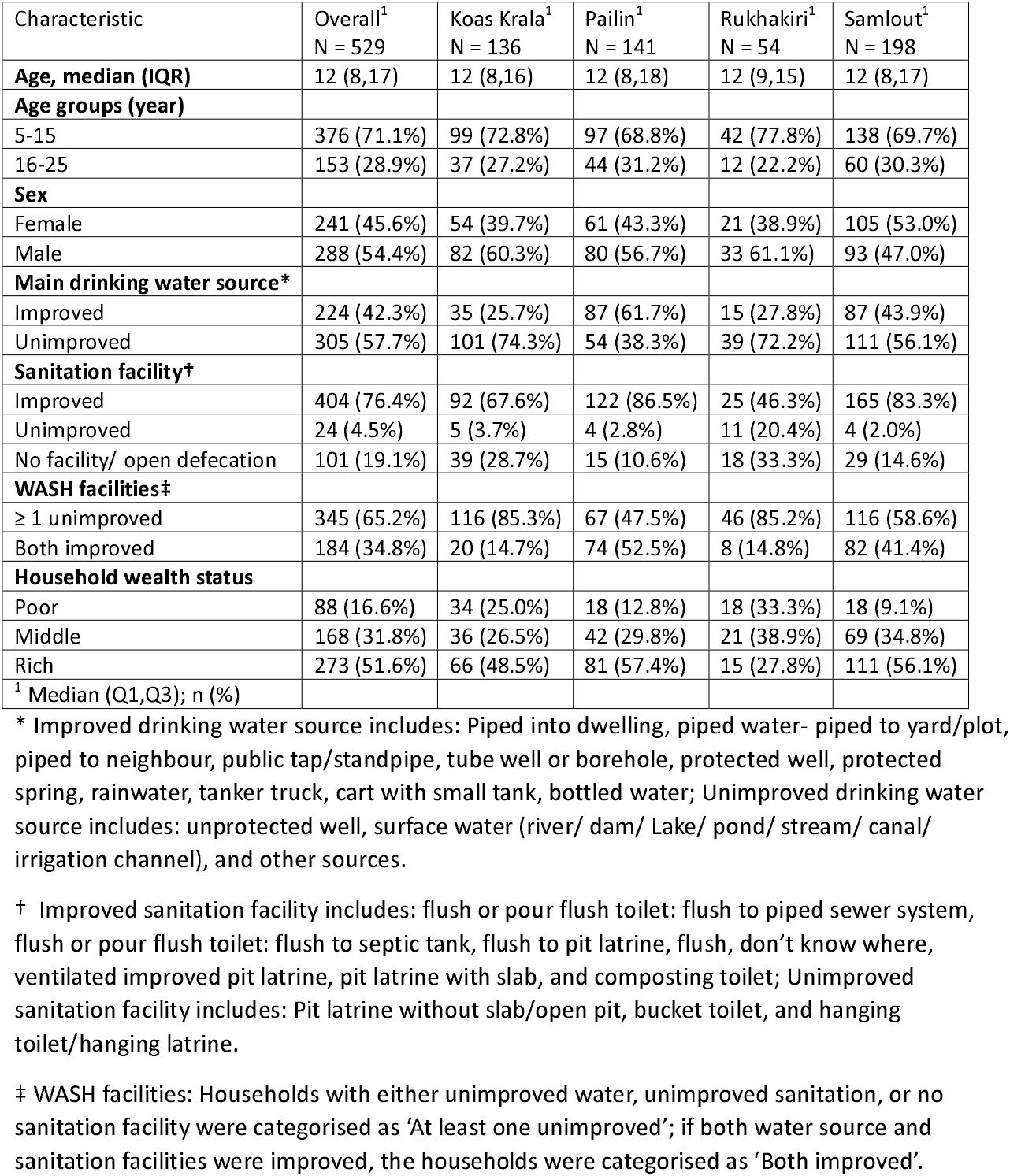
Overview of the demographic characteristics of participants aged 5-25 years included in the enteric fever seroincidence analysis in Cambodia, by district.

| Characteristic | Overall<sup>1</sup><br>N = 529 | Koas Krala<sup>1</sup><br>N = 136 | Pailin<sup>1</sup><br>N = 141 | Rukhakiri<sup>1</sup><br>N = 54 | Samlout<sup>1</sup><br>N = 198 |
| --- | --- | --- | --- | --- | --- |
| **Age, median (IQR)** | 12 (8,17) | 12 (8,16) | 12 (8,18) | 12 (9,15) | 12 (8,17) |
| **Age groups (year)** |  |  |  |  |  |
| 5-15 | 376 (71.1%) | 99 (72.8%) | 97 (68.8%) | 42 (77.8%) | 138 (69.7%) |
| 16-25 | 153 (28.9%) | 37 (27.2%) | 44 (31.2%) | 12 (22.2%) | 60 (30.3%) |
| **Sex** |  |  |  |  |  |
| Female | 241 (45.6%) | 54 (39.7%) | 61 (43.3%) | 21 (38.9%) | 105 (53.0%) |
| Male | 288 (54.4%) | 82 (60.3%) | 80 (56.7%) | 33 (61.1%) | 93 (47.0%) |
| **Main drinking water source\*** |  |  |  |  |  |
| Improved | 224 (42.3%) | 35 (25.7%) | 87 (61.7%) | 15 (27.8%) | 87 (43.9%) |
| Unimproved | 305 (57.7%) | 101 (74.3%) | 54 (38.3%) | 39 (72.2%) | 111 (56.1%) |
| **Sanitation facility†** |  |  |  |  |  |
| Improved | 404 (76.4%) | 92 (67.6%) | 122 (86.5%) | 25 (46.3%) | 165 (83.3%) |
| Unimproved | 24 (4.5%) | 5 (3.7%) | 4 (2.8%) | 11 (20.4%) | 4 (2.0%) |
| No facility/ open defecation | 101 (19.1%) | 39 (28.7%) | 15 (10.6%) | 18 (33.3%) | 29 (14.6%) |
| **WASH facilities‡** |  |  |  |  |  |
| ≥ 1 unimproved | 345 (65.2%) | 116 (85.3%) | 67 (47.5%) | 46 (85.2%) | 116 (58.6%) |
| Both improved | 184 (34.8%) | 20 (14.7%) | 74 (52.5%) | 8 (14.8%) | 82 (41.4%) |
| **Household wealth status** |  |  |  |  |  |
| Poor | 88 (16.6%) | 34 (25.0%) | 18 (12.8%) | 18 (33.3%) | 18 (9.1%) |
| Middle | 168 (31.8%) | 36 (26.5%) | 42 (29.8%) | 21 (38.9%) | 69 (34.8%) |
| Rich | 273 (51.6%) | 66 (48.5%) | 81 (57.4%) | 15 (27.8%) | 111 (56.1%) |
| <sup>1</sup> Median (Q1,Q3); n (%) |  |  |  |  |  |

\* Improved drinking water source includes: Piped into dwelling, piped water- piped to yard/plot, piped to neighbour, public tap/standpipe, tube well or borehole, protected well, protected spring, rainwater, tanker truck, cart with small tank, bottled water; Unimproved drinking water source includes: unprotected well, surface water (river/ dam/ Lake/ pond/ stream/ canal/ irrigation channel), and other sources.
† Improved sanitation facility includes: flush or pour flush toilet: flush to piped sewer system, flush or pour flush toilet: flush to septic tank, flush to pit latrine, flush, don't know where, ventilated improved pit latrine, pit latrine with slab, and composting toilet; Unimproved sanitation facility includes: Pit latrine without slab/open pit, bucket toilet, and hanging toilet/hanging latrine.
‡ WASH facilities: Households with either unimproved water, unimproved sanitation, or no sanitation facility were categorised as 'At least one unimproved'; if both water source and sanitation facilities were improved, the households were categorised as 'Both improved'.

**Figure 1.**
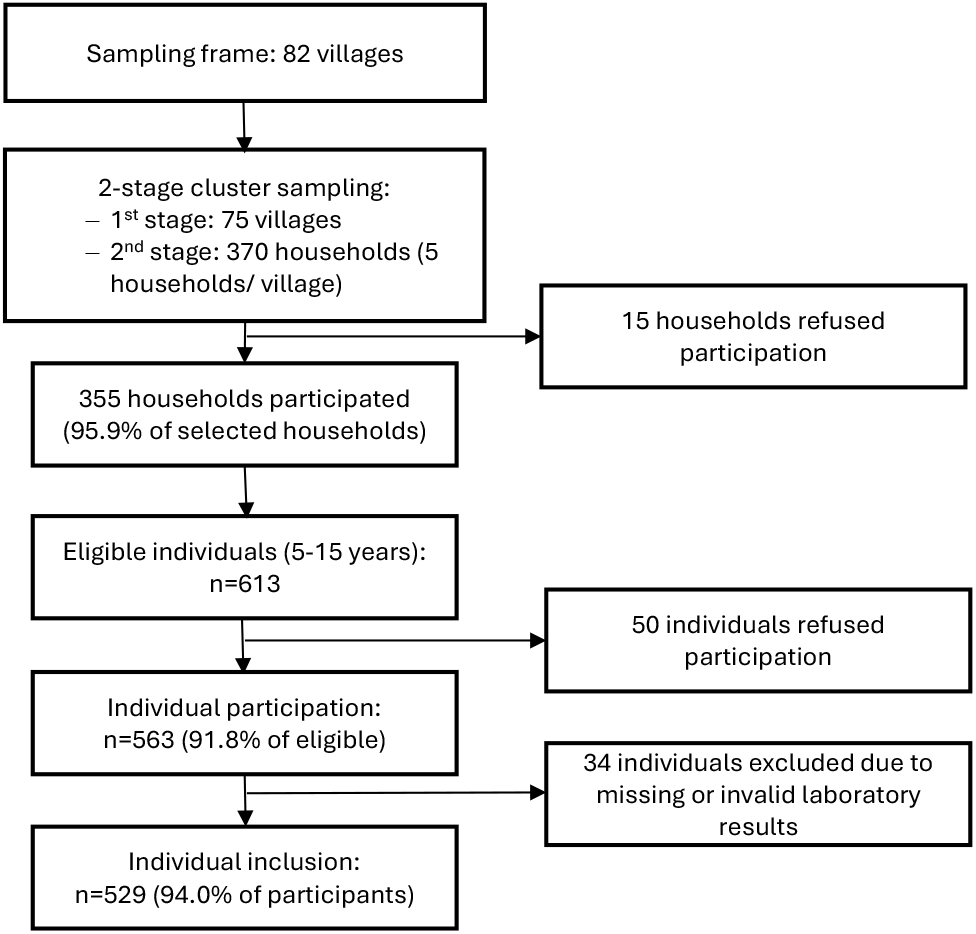
Flowchart of participant inclusion for the enteric fever seroincidence analysis. Villages and households were selected using a two-stage cluster sampling design. Of 370 selected households, 355 (95·9%) participated. Among 613 eligible individuals aged 5-25 years, 563 (91·8%) participated. After exclusion of participants with missing or invalid laboratory results, 529 (94.0%) individuals from 273 households were included in the analysis.

Quantitative antibody responses showed age-related patterns. Across the study population, IgA remained relatively stable across ages, and IgG increased with age (Figure 2). However, patterns varied by district. Koas Krala had both IgA and IgG declining with age, with IgG responses consistently higher across most ages. In Rukhakiri and Pailin, both IgA and IgG increased with age, while in Samlout, IgG increased but IgA declined. Pailin also showed higher IgA responses among older individuals.

**Figure 2.**
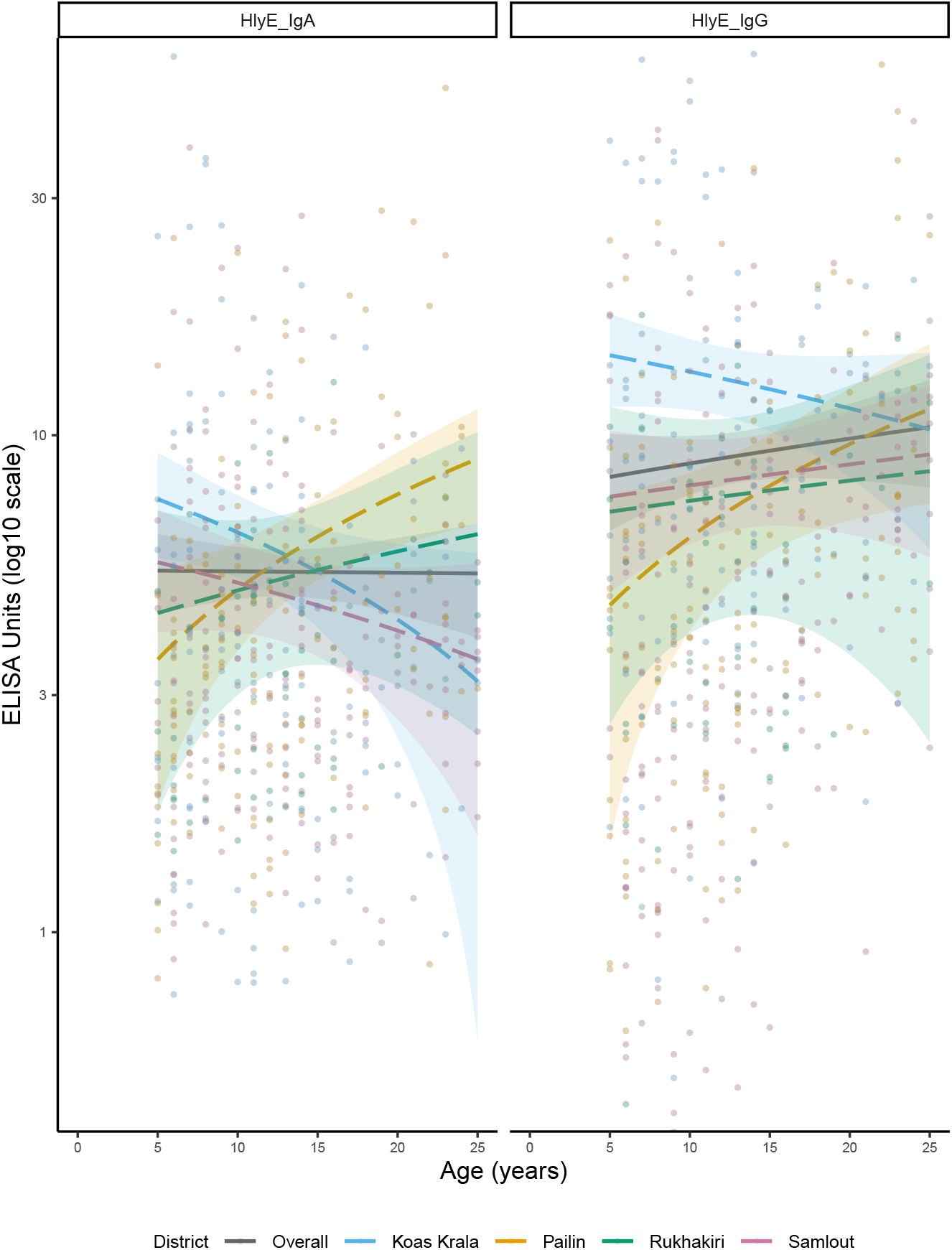
Age-dependent hemolysin E (HlyE) antigen of *Salmonella enterica*, IgA and IgG ELISA units, overall and by districts. Curves were fitted using generalised additive models (GAM). ELISA values are plotted on a log_10_ scale to better capture variation across antibody levels. The colored band along each curve indicated 95% confidence intervals. For visual clarity and interpretability, only values between 0.5 and 50 are shown, which cover the extended interquartile range. The median IgA ELISA value was 3.5 (IQR: 2.2–5.6; range: 0.006–76.5), and the median IgG value was 5.5 (IQR: 2.9–5.5; range: 0.01–112.0).

The overall combined IgA and IgG seroincidence of enteric fever per 1,000 person-years was 161.8 (95%CI: 144.6, 181.0) (Figure 3A). District-specific seroincidence was highest in Koas Krala (209.4, 95%CI: 158.6, 276.3), followed by Rukhakiri (161.5, 95%CI: 126.9, 205.5), Pailin (158. 5, 95%CI: 134.1, 187.3), and Samlout (139.7, 95%CI: 117.1, 276.3). Seroincidence was significantly higher in Koas Krala than in Samlout (p=0·03), while differences between other districts were not statistically significant (Supplementary material Table 2).

**Figure 3.**
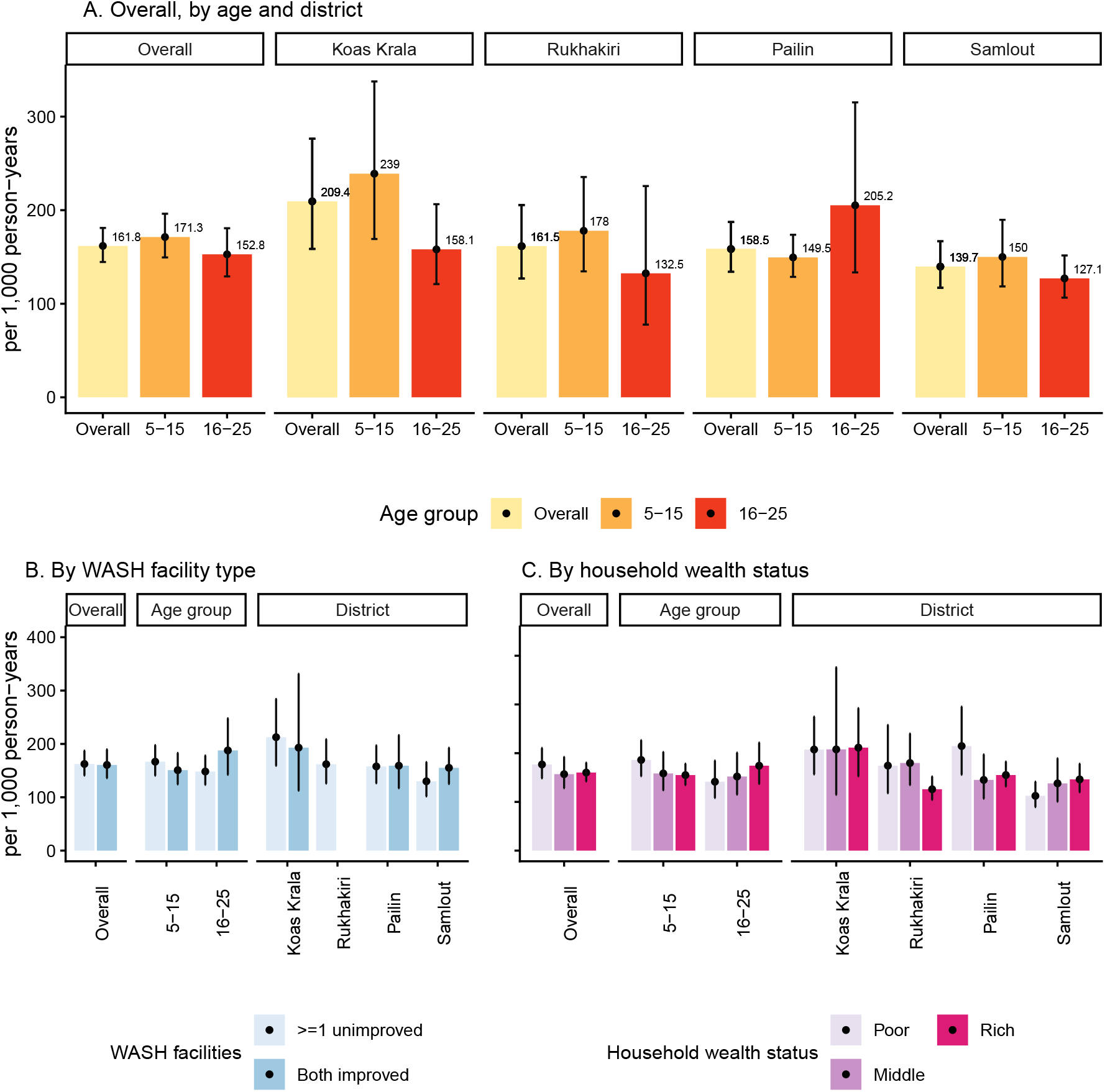
Enteric fever seroincidence estimates among participants aged 5-25 years from a cross-sectional serosurvey conducted in rural areas of western Cambodia based on quantitative antibody responses to HLyE IgA and IgG. (A) Overall seroincidence and estimates stratified by age group and district. (B) Seroincidence by household WASH facility type, overall and stratified by age group and district. (C) Seroincidence by household wealth status, overall and stratified by age group and district. Seroincidence estimates for participants from Rukhakiri households with improved WASH facilities are not shown due to a small sample size (n = 8).

Overall, children aged 5-15 years had a slightly higher seroincidence (171.3, 95%CI: 149.5, 196.2) than young adults aged 16-25 years (152.8, 95%CI: 129.2, 180.7), though not statistically significant (Figure 3A, Supplementary material Table 2). Age-related patterns varied by district. In Koas Krala, Rukhakiri, and Samlout, children had a higher seroincidence than older individuals, whereas the older individuals had a higher seroincidence than children in Pailin. However, no statistically significant age-group differences were observed within each district. Among 5-15-year-olds, seroincidence ranged from 149.5 (95%CI: 128.7, 173.7) in Pailin to 239.0 (95%CI:169.2, 337.6) in Koas Krala.

The overall seroincidence among all participants did not differ significantly by WASH access (both improved: 160.6, 95%CI: 136.0, 189.7 vs ≥1 unimproved: 162.4, 95%CI: 140.8, 187.4) (Figure3B, Supplementary material Table 2, Supplementary material Figure 1). Seroincidence was higher among participants from poorer households (194.0, 95% CI: 160.7, 234.0) than those from middle (169.2, 95% CI: 136.7, 209.5) or richer households (173.0, 95% CI: 152.6, 196.2) (Figure 3C), but the difference was not statistically significant (Supplementary material Table 2). Among children aged 5-15 years, seroincidence was slightly higher in those from households with ≥1 unimproved WASH (166.7, 95%CI: 140.6, 197.7 vs both improved facilities 150.8, 95%CI: 124.3, 183.0) and decreased with greater household wealth (poor: 207.5, 95% CI: 167.8, 256.5; middle: 174.1, 95% CI: 134.6, 225.2; rich: 170.2, 95% CI: 147.1, 197.0). However, the differences were not statistically significant (Supplementary material Table 2).

Monthly seroincidence was consistent at around 150/1,000 person-years, except for a low in June (111.2/1,000 person years, 95%CI: 90.8, 136.2), and a high in August (331.1/1,000 person years, 95%CI: 244.4, 448.6). Seroincidence in August and July differed significantly from other months (August: p<0.001; July: p<0.05) (Supplementary material Table 2, Supplementary material Table 3).

## Discussion

In this population-based, representative serosurvey in rural, Western Cambodia, we observed a high enteric fever (*S*. Typhi and *S*. Paratyphi infection) seroincidence rate of 161.8/1,000 person-years that indicating a substantially higher burden than suggested by previously reported clinical incidence.

The Global Burden of Disease (GBD) study estimated a clinical incidence, distinct from seroincidence, of enteric fever at 1.1/ 1,000 person-years among individuals under 20 in Cambodia.^2^ Because no Cambodia-specific seroincidence-to-clinical incidence conversion factors are available, we contextualised our findings by applying conversion factors derived from prior studies using the same seroincidence estimation methods.^8,13^ These studies reported that the seroincidence rate was 16-32 times higher than prospective blood culture surveillance data, after adjustment for blood culture sensitivity (59%) and probability of health-seeking behaviour, in Bangladesh, Pakistan and Nepal. Applying this seroincidence-to-clinical incidence ratio, our findings suggest a clinical incidence of enteric fever in Cambodia may be 5.1-10.1/1,000 person-years, which is 5-10 times higher than GBD estimates. Such discrepancies are expected, as facility-based passive surveillance typically underestimates clinical incidence due to variation in care-seeking behaviour, limited diagnostic sensitivity, antibiotic use before culture, and failure to capture mild or subclinical cases. Furthermore, available data are largely from urban settings, with little from rural areas.^3,14^ In addition, GBD estimates for Cambodia relied on modelling based on covariates such as education and WASH, and incidence data from neighbouring countries, and regional and global patterns, because enteric fever-specific national data were lacking.^2^ Our findings highlight the value of seroincidence data, derived from population-based surveillance, in providing a more complete picture of infection occurrence in resource-limited settings.

Our findings align with the existing understanding of a high burden of enteric fever among children in Cambodia, meeting the definition of a high-incidence typhoid fever setting (>1/1,000 case-years).^1,15^ In our study, seroincidence among children aged 5-15 years ranged from 149.5 to 239.0/ 1,000 person-years across districts. Although direct estimates of enteric fever incidence in this age group are unavailable, a hospital-based study in Siem Reap, a neighbouring province, reported typhoid fever incidence, according to blood culture or discharge diagnosis, between 0·6 and 11·4/1,000 person-years.^16^ Limited existing data from hospital-based studies in Siem Reap (2007-2016) suggest *S*. Typhi accounts for 40%-90% of enteric fever cases among children under 15.^16,17^ Applying this attribution range and the same seroincidence-to-clinical incidence ratio (16-32) yields inferred *S*. Typhi incidence of 1.8-13.4 /1,000 person-years.^8,13^ These estimates are higher but overlap with prior observations and suggest Cambodia’s classification as a high-incidence typhoid fever setting.^15^ Our seroincidence estimates confirmed a high enteric fever burden among children and argue for consideration of TCV as part of Cambodia’s public health strategy.

We observed district-level differences in age-specific seroincidence and age-dependent quantitative ELISA responses, reflecting spatial heterogeneity in enteric fever transmission. In Koas Krala, and to a lesser extent Rukhakiri and Samlout, all had higher seroincidence among children aged 5-15 years than older individuals, consistent with the known higher global burden among younger children.^1^ These patterns aligned with age-related antibody levels, with declining IgA with age in Koas Krola and Samlout, a modest increase in Rukhakiri, while a declining IgG with age in Koas Krala. In contrast, Pailin showed higher seroincidence among individuals aged 16-25 years. Although the cause remains unclear, one possible explanation is the age-, location-, and time-specific variations in the distribution of *Salmonalla serovars*, and with a greater contribution of *S*. Paratyphi to adult enteric fever in our setting. This interpretation is supported by a higher quantitative IgA response among adults in Pailin.^18^ Evidence from previous studies further supports this hypothesis. As mentioned above, *S*. Typhi accounted for 40-90% of enteric fever cases among children under 15 in Siem Reap from 2007-2016.^16,17^ In contrast, in Phnom Penh, Cambodia’s capital, a study conducted in a hospital where most patients were adults, showed shifted serovar distribution: *S*. Typhi predominated during 2008-2012 (74%, n=35), followed by a sharp rise in *S*. Paratyphi in 2013 (82%, n=72) while *S*. Typhi case numbers stayed relatively stable.^19^ During 2014-2015, *S*. Paratyphi persistently accounted for 80-90% of adult cases. A high burden of *S*. Paratyphi among adults could therefore explain both the elevated seroincidence in the 16-25-year age group and the high overall seroincidence. These findings highlight the importance of investigating local epidemiological data to shape context-specific intervention strategies.

Using our seroincidence estimates, we were able to identify priority populations for targeted interventions in our study areas, including children aged 5-15 years in high-incidence districts (Koas Krala and Rukhakiri) and from households with poorer WASH access and lower wealth status. Although differences by household WASH access and wealth status were not statistically significant, seroincidence among children aged 5-15 years was numerically higher in those from households with poor WASH access and lower wealth status. The two highest-seroincidence districts, Koas Krala and Rukhakiri, also had the poorest WASH access, and a higher proportion of participants lived in ‘Poor’ households, suggesting these factors may contribute to increased transmission. In contrast, among individuals 16-25 years, numerically higher seroincidence was observed among those from households with both improved WASH and higher wealth status. This may reflect the association between enteric fever and the unmeasured factors among adults, such as behavioural or occupational exposures. While interventions to improve WASH should be prioritised in high-incidence districts to reduce both *S*. Typhi and *S*. Paratyphi infections, future studies should investigate other potential determinants related to adults to inform context-specific risk-reduction strategies.

This study has several limitations. First, we were unable to include children aged under five years, who bear the greatest global burden of enteric fever.^1^ Although dried blood spot (DBS) samples were collected from this age group, prolonged ambient-temperature storage raised concern about antibody degradation. Therefore, these samples were excluded from this study. Second, the serological assay cannot distinguish between *S*. Typhi and *S*. Paratyphi infections. As TCV only protects against *S*. Typhi, this limits the specificity of our findings for vaccine impact assessment. Additional blood culture data would be useful to inform the relative contribution of *S*. Typhi versus *S*. Paratyphi, which would improve the interpretation of seroincidence estimates and strengthen evidence to inform TVC recommendations. Third, the seroincidence-clinical incidence ratio applied here for the comparison with previous estimates on clinical incidence, derives from other countries and may not fully represent the Cambodian context due to variations in healthcare access.

In addition, the seroincidence model assumes a constant force of infection over time and does not account for potential seasonal variation in transmission. In Cambodia, enteric fever typically peaks during the rainy season (May to October),^16^ and our eight-month blood sample collection period included five months of this peak, potentially inflating annual seroincidence estimates. The August peak coincided with data collection in Koas Krala, the highest-seroincidence district, where approximately half of the participants (n=67) were recruited in August. In Koas Krala, quantitative IgG were also high across all ages, suggesting sampling may have occurred during a period of heightened transmission. Future studies should collect samples across all districts, ideally for multiple years, to determine the impact of seasonal variability on seroincidence estimation.

Additionally, the study site may not fully represent the national context, as regional variations in WASH infrastructures, healthcare access, and the relative attributions of *S*. Typhi and *S*. Paratyphi could influence enteric fever incidence. However, as discussed previously, the overlap between our estimates among children under 15 and those reported from Siem Reap suggests that our findings from this age group may be generalisable to rural western Cambodia. Further studies in different contexts, particularly in urban settings, are needed to strengthen the evidence base for TCV introduction across diverse settings in Cambodia.

Our study provides a community-based estimate of enteric fever seroincidence in rural areas in western Cambodia, indicating a substantially higher disease burden than previously reported. These findings contribute evidence to ongoing discussions on TCV introduction in Cambodia. More broadly, this study demonstrates the utility of serosurveillance as a valuable tool that overcomes underreporting and underdiagnosis in passive surveillance systems. By leveraging antibody kinetics and quantitative serological assays, this approach for seroincidence estimates could be expanded to other infectious diseases, offering a scalable method for disease surveillance in resource-limited settings, where traditional facility-based surveillance may be inadequate or unavailable.

## Funding

This research was funded in whole, or in part, by the Wellcome Trust [215604/Z/19/Z], National Institutes of Health Fogarty International Centre (K01TW012177), and PATH. For the purpose of Open Access, the author has applied a CC BY public copyright license to any Author Accepted Manuscript version arising from this submission. The funders of the study had no role in the study design, data collection, data analysis, data interpretation, or writing of the report.

## Contributors

MZ, EB, SJL, RJM, TJP, NPJD, RC, YL, and KA conceptualised the study. EB, RJM, TJP, NPJD, YL, and KA acquired the funding, MZ, TP, EB, RC, KA developed the methodology. MZ, RT, LD, PC, and VM supervised enrolment and data collection. NS, NC, and RC supervised laboratory work. MZ and KA did the data analysis and data visualisation. MZ and KA wrote the original draft of the manuscript. All authors reviewed and edited the manuscript. KA accessed and verified the data. All authors had full access to all the data in the study and had final responsibility for the decision to submit for publication.

## Declaration of interests

We declare no competing interests.

## Data sharing

Data collected for the study, including de-identified individual participant data, will be made available to others. These data will be available with publication via the MORU data sharing committee. Access will be provided for analysis by bona fide researchers with or without investigator support, after approval of a proposal, and upon a signed data access agreement.

## Acknowledgments

This work was supported by Wellcome Trust [215604/Z/19/Z], National Institutes of Health Fogarty International Center (K01TW012177), and PATH. We would like to acknowledge the contributions of the field teams in Cambodia and the support teams at the Mahidol-Oxford Tropical Medicine Research Unit for their essential roles in data and sample collection, study monitoring, and laboratory procedures.

Declaration of generative AI and AI-assisted technologies in the manuscript preparation process During the preparation of this work the authors used ChatGPT 5.1 to assist copy-editing. After using this tool, the authors reviewed and edited the content as needed and take full responsibility for the content of the published article.

## Notes

### Competing Interest Statement

The authors have declared no competing interest.

### Clinical Protocols

https://doi.org/10.1136/bmjopen-2023-081079

### Author Declarations

This study protocol was approved by the Oxford Tropical Research Ethics Committee (OxTREC Ref: 6-22) and the Cambodian National Ethics Committee for Health Research (December 23, 2022 NECHR).

## References

1. Murthy S, Hagedoorn NN, Faigan S, et al. Global typhoid fever incidence: an updated systematic review with meta-analysis. Lancet Infect Dis 2025; 0. DOI:10.1016/S1473-3099(25)00359-7.

2. Institute for Health Metrics and Evaluation (IHME). GBD Results. Seattle, WA: IHME, University of Washington, 2025 https://vizhub.healthdata.org/gbd-results/ (accessed Nov 25, 2025).

3. Antillon M, Saad NJ, Baker S, Pollard AJ, Pitzer VE. The Relationship Between Blood Sample Volume and Diagnostic Sensitivity of Blood Culture for Typhoid and Paratyphoid Fever: A Systematic Review and Meta-Analysis. J Infect Dis 2018; 218: S255–67.

4. Andrews JR, Baker S, Marks F, et al. Typhoid conjugate vaccines: a new tool in the fight against antimicrobial resistance. Lancet Infect Dis 2019; 19: e26–30.

5. World Health Organization. Typhoid vaccines: WHO position paper. 2018; published online March. https://www.who.int/publications/i/item/whio-wer9313 (accessed Dec 15, 2024).

6. Chandna A, Chew R, Shwe Nwe Htun N, et al. Defining the burden of febrile illness in rural South and Southeast Asia: an open letter to announce the launch of the Rural Febrile Illness project. Wellcome Open Res 2022; 6: 64.

7. Zhang M, Htun NSN, Islam S, et al. Defining the hidden burden of disease in rural communities in Bangladesh, Cambodia and Thailand: a cross-sectional household health survey protocol. BMJ Open 2024; 14: e081079.

8. Aiemjoy K, Seidman JC, Saha S, et al. Estimating typhoid incidence from community-based serosurveys: a multicohort study. Lancet Microbe 2022; 3: e578–87.

9. Croft, Trevor N., Aileen M. J. Marshall, Courtney K. Allen, et al. Guide to DHS Statistics. Rockville, Maryland, USA: ICF., 2018.

10. Metrics for Management. The Cambodia Equity Tool. 2016; published online Nov 1. https://www.equitytool.org/cambodia/ (accessed April 30, 2024).

11. Wood SN. Generalized Additive Models: An Introduction with R, Second Edition, 2nd edition. Chapman and Hall/CRC, 2017.

12. Lai KW, Orwa C, Seidman JC, et al. serocalculator, an R package for estimating seroincidence from cross-sectional serological data. 2025; : 2025.06.04.25328941.

13. Garrett DO, Longley AT, Aiemjoy K, et al. Incidence of typhoid and paratyphoid fever in Bangladesh, Nepal, and Pakistan: results of the Surveillance for Enteric Fever in Asia Project. Lancet Glob Health 2022; 10: e978–88.

14. Garrett D. The surveillance for enteric fever in Asia project (SEAP): Estimating the community burden of enteric fever. Int J Infect Dis 2016; 45: 64.

15. Crump JA, Luby SP, Mintz ED. The global burden of typhoid fever. Bull World Health Organ 2004; 82: 346–53.

16. Thanh DP, Thompson CN, Rabaa MA, et al. The Molecular and Spatial Epidemiology of Typhoid Fever in Rural Cambodia. PLoS Negl Trop Dis 2016; 10: e0004785.

17. Kheng C, Meas V, Pen S, Sar P, Turner P. Salmonella Typhi and Paratyphi A infections in Cambodian children, 2012–2016. Int J Infect Dis 2020; 97: 334–6.

18. Seidman JC, Aiemjoy K, Adnan M, et al. Evaluating the accuracy of Salmonella Typhi Hemolysin E and lipopolysaccharide IgA to discriminate enteric fever from other febrile illnesses in South Asia. 2025; : 2025.06.20.25329792.

19. Kuijpers LMF, Phe T, Veng CH, et al. The clinical and microbiological characteristics of enteric fever in Cambodia, 2008-2015. PLoS Negl Trop Dis 2017; 11: e0005964.

