## Supplementary materials for "Estimating enteric fever seroincidence in rural western Cambodia: findings from a population-based cross-sectional serosurvey"

**Table 1. Seroincidence estimated by IgA or IgG alone, overall, by age group and district.**

| Characteristics | Overall* | Koas Krala* | Pailin* | Rukhakiri* | Samlout* |
|---|---|---|---|---|---|
| **IgA** | | | | | |
| Overall | 156.5 (132.2, 185.3) | 154.4 (105.2, 226.7) | 174.5 (130.2, 233.9) | 134.8 (83.5, 217.5) | 152.3 (116.7, 198.8) |
| 5-15 years | 167.6 (138.7, 202.6) | 178 (110, 288.2) | 153.9 (117.3, 201.8) | 132.5 (76.4, 229.9) | 182.7 (137.2, 243.3) |
| 16-25 years | 127.7 (97, 168.3) | 99.8 (68.2, 146.1) | 275.9 (110.1, 691.8) | 144.6 (68.8, 304.1) | 97.4 (71.2, 133.4) |
| **IgG** | | | | | |
| Overall | 165.2 (145.8, 187.2) | 257 (201, 328.6) | 149.8 (126.5, 177.4) | 180.4 (153.8, 211.6) | 132.5 (107.8, 162.9) |
| 5-15 years | 157.7 (135.7, 183.3) | 258.7 (194, 345) | 134.2 (111.7, 161.3) | 194.7 (163.7, 231.6) | 120.6 (92.7, 156.7) |
| 16-25 years | 189.8 (162, 222.5) | 251 (193.3, 326.1) | 201.7 (143.9, 282.7) | 135.3 (86, 212.7) | 168.1 (136.1, 207.7) |
| *Seroincidence (95%CI), all reported per 1,000 person-years | | | | | |

**Table 2. Comparisons of enteric fever seroincidence using two-sample z-test, by district, age group, WASH access, household wealth status, and month of data collection.**

| Variable | Comparison group | Seroincidence differences (95%CI)* | z statistic | p-value |
|---|---|---|---|---|
| **District** | | | | |
| | Koas Krala vs.Samlout | 69.6 (6.5, 132.7) | 2.1619354 | 0.030623 |
| | Koas Krala vs.Pailin | 50.9 (-13, 114.7) | 1.5619949 | 0.118289 |
| | Koas Krala vs.Rukhakiri | 47.9 (-22, 117.8) | 1.3427298 | 0.17936 |
| | Pailin vs.Rukhakiri | -3 (-50.1, 44.1) | -0.1242687 | 0.901102 |
| | Pailin vs.Samlout | 18.7 (-17.5, 54.9) | 1.0141354 | 0.310518 |
| | Rukhakiri vs.Samlout | 21.7 (-24.4, 67.8) | 0.9238461 | 0.355566 |
| **Age group** | | | | |
| | 16-25 years vs. 5-15 years | -18.5 (-53.1, 16.1) | -1.0494292 | 0.293981 |
| Age group by district | | | | |
| Koas Krala | 16-25 years vs. 5-15 years | -80.9 (-173.7, 11.8) | -1.7109965 | 0.087082 |
| Rukhakiri | 16-25 years vs. 5-15 years | -45.5 (-131.9, 40.9) | -1.032052 | 0.302048 |
| Pailin | 16-25 years vs. 5-15 years | 55.6 (-35.3, 146.5) | 1.1995785 | 0.230303 |
| Samlout | 16-25 years vs. 5-15 years | -22.9 (-64.7, 18.9) | -1.0732852 | 0.283143 |
| **WASH access** | | | | |
| | ≥1 unimproved vs. Both improved | 1.8 (-33.6, 37.2) | 0.099512924 | 0.920731 |
| WASH access by district | | | | |

| | | | | |
|---|---|---|---|---|
| Koas Krala | ≥1 unimproved vs. Both improved | 19.6 (-101.6, 140.8) | 0.317213171 | 0.751082 |
| Rukhakiri | ≥1 unimproved vs. Both improved | 3.8 (-60.3, 67.9) | 0.117164165 | 0.90673 |
| Pailin | ≥1 unimproved vs. Both improved | -1.3 (-61.6, 58.9) | -0.043359932 | 0.965415 |
| Samlout | ≥1 unimproved vs. Both improved | -25.3 (-71.6, 21) | -1.071436529 | 0.283973 |
| WASH access by age group | | | | |
| 16-25 years | ≥1 unimproved vs. Both improved | -39.5 (-98.4, 19.5) | -1.312910832 | 0.189213 |
| 5-15 years | ≥1 unimproved vs. Both improved | 15.9 (-24.8, 56.6) | 0.766736319 | 0.443238 |
| **Household wealth status** | | | | |
| | Poor vs. Middle | 19.9 (-24.4, 64.2) | 0.88027989 | 0.378708 |
| | Poor vs. Rich | 16.7 (-19.7, 53.1) | 0.898276189 | 0.369038 |
| | Middle vs. Rich | -3.2 (-40.3, 33.8) | -0.170972841 | 0.864245 |
| Household wealth status by district | | | | |
| Koas Krala | Poor vs. Middle | -0.3 (-136.9, 136.2) | -0.004530605 | 0.996385 |
| | Poor vs. Rich | -3.9 (-94, 86.2) | -0.085042552 | 0.932228 |
| | Middle vs. Rich | -3.6 (-144.8, 137.6) | -0.049876798 | 0.960221 |
| Rukhakiri | Poor vs. Middle | -5.5 (-91.2, 80.3) | -0.124873789 | 0.900623 |
| | Poor vs. Rich | 48.4 (-23.8, 120.6) | 1.314130348 | 0.188802 |
| | Middle vs. Rich | 53.9 (-3.3, 111.1) | 1.845607444 | 0.064949 |
| Pailin | Poor vs. Middle | 69.4 (-12.4, 151.3) | 1.663513065 | 0.09621 |
| | Poor vs. Rich | 59.4 (-13.7, 132.6) | 1.59269191 | 0.111229 |
| | Middle vs. Rich | -10 (-61.4, 41.4) | -0.381721413 | 0.702668 |
| Samlout | Poor vs. Middle | -25.6 (-76.3, 25.2) | -0.987813232 | 0.323244 |
| | Poor vs. Rich | -33.8 (-72.6, 5) | -1.709325066 | 0.087391 |
| | Middle vs. Rich | -8.2 (-60.7, 44.2) | -0.307196074 | 0.758694 |
| Household wealth status by age group | | | | |
| 16-25 years | Poor vs. Middle | -10.7 (-67.5, 46.1) | -0.369037443 | 0.7121 |
| | Poor vs. Rich | -32.9 (-89.3, 23.6) | -1.141076156 | 0.253838 |
| | Middle vs. Rich | -22.2 (-81.8, 37.4) | -0.729079617 | 0.465953 |
| 5-15 years | Poor vs. Middle | 27.7 (-25.6, 81) | 1.020037808 | 0.307711 |
| | Poor vs. Rich | 31.1 (-11.5, 73.6) | 1.429762508 | 0.152785 |
| | Middle vs. Rich | 3.3 (-41.1, 47.8) | 0.146484589 | 0.883539 |
| **Month of data collection** | | | | |
| | Apr, May vs. Aug | -180.5 (-284.1, -76.9) | -3.414646782 | 0.000639 |
| | Apr, May vs. Jun | 39.4 (5.7, 73.1) | 2.293753299 | 0.021805 |
| | Apr, May vs. Oct, Nov | 0.6 (-39.1, 40.4) | 0.030594313 | 0.975593 |
| | Apr, May vs. Sep | 2.1 (-31.9, 36.1) | 0.12044058 | 0.904134 |

| | | | | |
|---|---|---|---|---|
| | Aug vs. Jun | 219.9 (116.9, 322.9) | 4.183158951 | 2.87E-05 |
| | Aug vs. Oct, Nov | 181.1 (75.9, 286.3) | 3.374805416 | 0.000739 |
| | Aug vs. Sep | 182.6 (79.4, 285.7) | 3.46991203 | 0.000521 |
| | Jun vs. Oct, Nov | -38.8 (-77.1, -0.5) | -1.986599951 | 0.046967 |
| | Jun vs. Sep | -37.3 (-69.5, -5.1) | -2.2719975 | 0.023087 |
| | Oct, Nov vs. Sep | 1.5 (-37.1, 40) | 0.074601211 | 0.940532 |
| *Seroincidence differences (95%CI) reported per 1,000 person-years | | | | |

**Table 3. HLyE IgA and IgG seroincidence by month of data collection**

| **Data collection period *** | **Number of participants** | **Seroincidence (95%CI) / 1,000 person-years^** |
|---|---|---|
| Apr, May† | 137 | 150.7 (127.6, 177.9) |
| Jun | 70 | 111.2 (90.8, 136.2) |
| Aug | 96 | 331.1 (244.4, 448.6) |
| Sep | 118 | 148.6 (127.3, 173.4) |
| Oct, Nov† | 108 | 150.0 (122.1, 184.4) |

* Sample collection occurred during 8 months of the recruitment period (all activities paused in July due to local political activities, and no samples were collected in December). The data collection months of each district are: Rukhakiri- April (n=46), May (n=8); Koas Krala- August (n=67), September (n=69); Samlout- May (n=83), June (n=70), August (n=29), November (n=16); Pailin- September (n=49), October (n=92).

† Data from April and May were combined, as were the months October and November, due to the small number of participants recruited in April and November.

^Seroincidence differed significantly by month of data collection, with August showing higher estimates than all other months (p<0·001), and June differing significantly from April, May, September, and October, November (p<0·05).

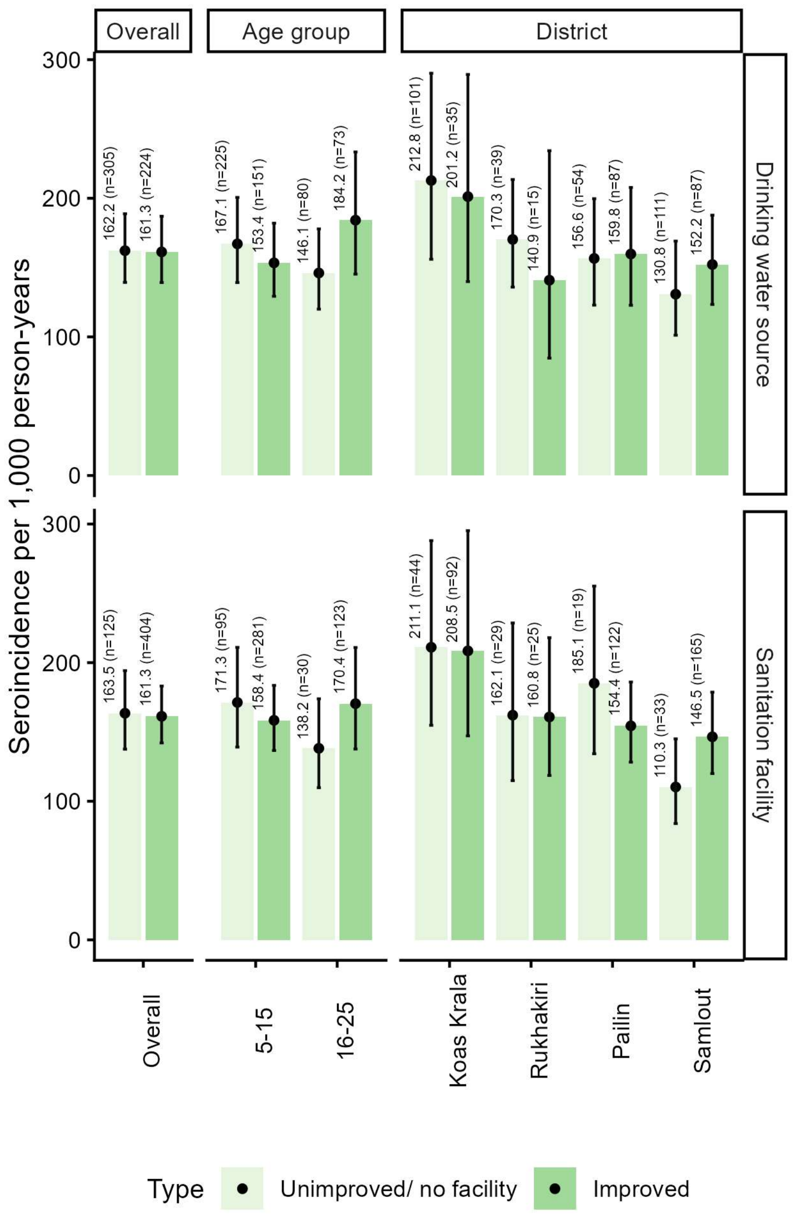


**Figure 1. Enteric fever seroincidence estimates based on quantitative antibody responses to HLyE IgA and IgG, by drinking water source (top) and sanitation facility type (bottom), overall and stratified by age and district.**

No statistically significant differences in seroincidence were observed between improved and unimproved drinking water sources or sanitation facilities across age groups, districts, or overall.

For sanitation facilities, the "Unimproved" and "No facilities" categories were combined due to small sample sizes. Seroincidence estimates are shown, with the number of observations indicated in parentheses.
